# Translational PBPK-QSP modeling platform for antibody-drug conjugates (ADC): within-target and cross-pathway validation to bridge preclinical and clinical results

**DOI:** 10.64898/2026.01.30.26345218

**Authors:** Andreas D. Meid, Ignacio Leiva-Escobar, Siak-Leng Choi, Delphine Valente

## Abstract

We designed a platform model that integrates physiologically-based pharmacokinetic (PBPK) modeling with quantitative systems pharmacology (QSP) to bridge translational challenges in antibody-drug conjugate (ADC) development. The PBPK-QSP platform model was developed for the ADC trastuzumab emtansine (T-DM1) in breast cancer patients. This mechanistic framework facilitates translation across preclinical in vitro experiments, in vivo studies, and clinical trials, supporting decision-making for novel ADCs. The PBPK-QSP model adequately predicts preclinical and clinical PK and PD data from two additional ADCs: trastuzumab deruxtecan (T-Dxd) and tusamitamab ravtansine. For within-target validation with T-Dxd in breast cancer, despite extensive preclinical calibration, efficacy predictions were initially overly optimistic compared to T-DM1 validation experience with the model and aggregated phase II trial data. Individual patient data from a phase II T-Dxd trial allowed evaluation of model performance and quantification of translational uncertainty in predicting clinical outcomes using preclinical experiments. Cross-pathway validation with tusamitamab ravtansine in non-small cell lung cancer has revealed the importance of incorporating a resistance module to describe clinical efficacy adequately. Clinical trial simulations for tusamitamab ravtansine subsequently inform that alternative fractional dosing could offer a potential efficacy advantage compared to existing clinical dosing. We integrated these insights into a practical recommended workflow for translational development programs, which addresses the key challenges in parameter estimation, data requirements, and uncertainty quantification in the key system parameters for each indication and cancer type. Ultimately, integrating an interactive modeling platform with a structured workflow to mitigate the risks of human translation and to potentially improve the clinical benefits of novel ADCs in oncology drug development.

## Introduction

Quantitative Systems Pharmacology (QSP) has emerged as a pivotal tool in model-informed drug development [1], effectively bridging the translational gap between preclinical findings and clinical outcomes [2-4]. Cucurull-Sanchez recently highlighted QSP’s transformative role in pharmaceutical decision-making, emphasizing its potential to enhance predictability and streamline development pipelines [5]. Numerous QSP models have supported the translational steps from in vitro experiments, in vivo animal studies, and their scale-up into clinical trials with patients [6], including those for antibody-drug conjugates (ADCs). QSP models typically fall into two categories: “fit-for-purpose” models tailored for specific projects, and “platform” models offering versatile, reusable and adaptable frameworks with a broader applicability across various projects [5, 7]. However, for platform models to provide meaningful value in clinical development, where translational barriers present significant challenges, they must reliably address critical aspects such as animal-to-human translation and a thorough understanding of pharmacokinetic/pharmacodynamic (PK/PD) relationships across the full translational spectrum—from in vitro experiments and in vivo animal studies to clinical trials with patients. Recent case studies with the ADC trastuzumab emtansine (T-DM1) have demonstrated how translational QSP models can effectively support drug development by addressing these challenges [8].

While QSP models are valuable, especially for internal decision-making within the industry, they are not without challenges: Larry Lesko has raised concerns regarding cost, complexity, and the uncertain regulatory landscape surrounding QSP models, suggesting that their adoption may sometimes be driven more by technological enthusiasm than by clear, practical need [9]. The standardized and potentially more efficient approach of a platform model approach could address some of these concerns. However, model validation remains as a substantial challenge, but is crucial to ensure the reliability, relevance, and predictive power of QSP models prior to using them for prospective simulation to inform decisions. A key obstacle is often availability of external clinical data sets which can be difficult or impossible to obtain for novel drugs in development. Furthermore, Androulakis emphasized the need for comprehensive validation strategies that address the underlying biological complexity, proposing a spectrum of within-target to cross-pathway validation approaches [10].

Our work builds upon these principles to develop and validate an ADC platform model with practical translational utility. Initially developed using T-DM1 as a foundation, it leverages established physiologically-based pharmacokinetic (PBPK) translation from mouse to human and PD/efficacy translation from in vitro/in vivo preclinical to clinical settings. We subsequently applied this platform in a within-target validation to trastuzumab deruxtecan (T-Dxd), using phase II individual patient data (IPD) [11]. We then expanded to cross-pathway validation with an investigational ADC targeting CEACAM5 (tusamitamab ravtansine) [12-14], mimicking real-world application scenarios in drug development. This comprehensive approach addresses the critical need for interactive translational platforms that can predict PK, PD, and efficacy for novel cytotoxic ADCs in cancer patients. Beyond, this article also provides practical guidelines for utility of this approach - including data requirements, parameter sensitivity considerations, and strategies for managing uncertainty. It represents a balanced approach to QSP modeling that aligns with industry needs while addressing key concerns around complexity, uncertainty, and validation, ultimately enhancing model-informed decision-making in ADC development.

## Methods

Initially, we derived a platform model based on a PBPK model for T-DM1and connected it with a tumoral disposition model to reach single cells which pharmacodynamically linked it to tumor growth inhibition. Thus, the molecular interactions of the antibody part (mAb) and cytotoxic payload at the cellular level are key elements of this PBPK-QSP model **(Supplementary Figure S1**). Once this model has been qualified, it was used for simulations and validation studies for new ADCs, including within-target (T-Dxd) and the cross-pathway (tusamitamab ravtansine, SAR408701) (**Figure 1**). The following sections outline the steps involved in developing the platform model, the methodologies used for validation and performance assessment, and the necessary steps for adapting such models to new substances.

**Figure 1.**
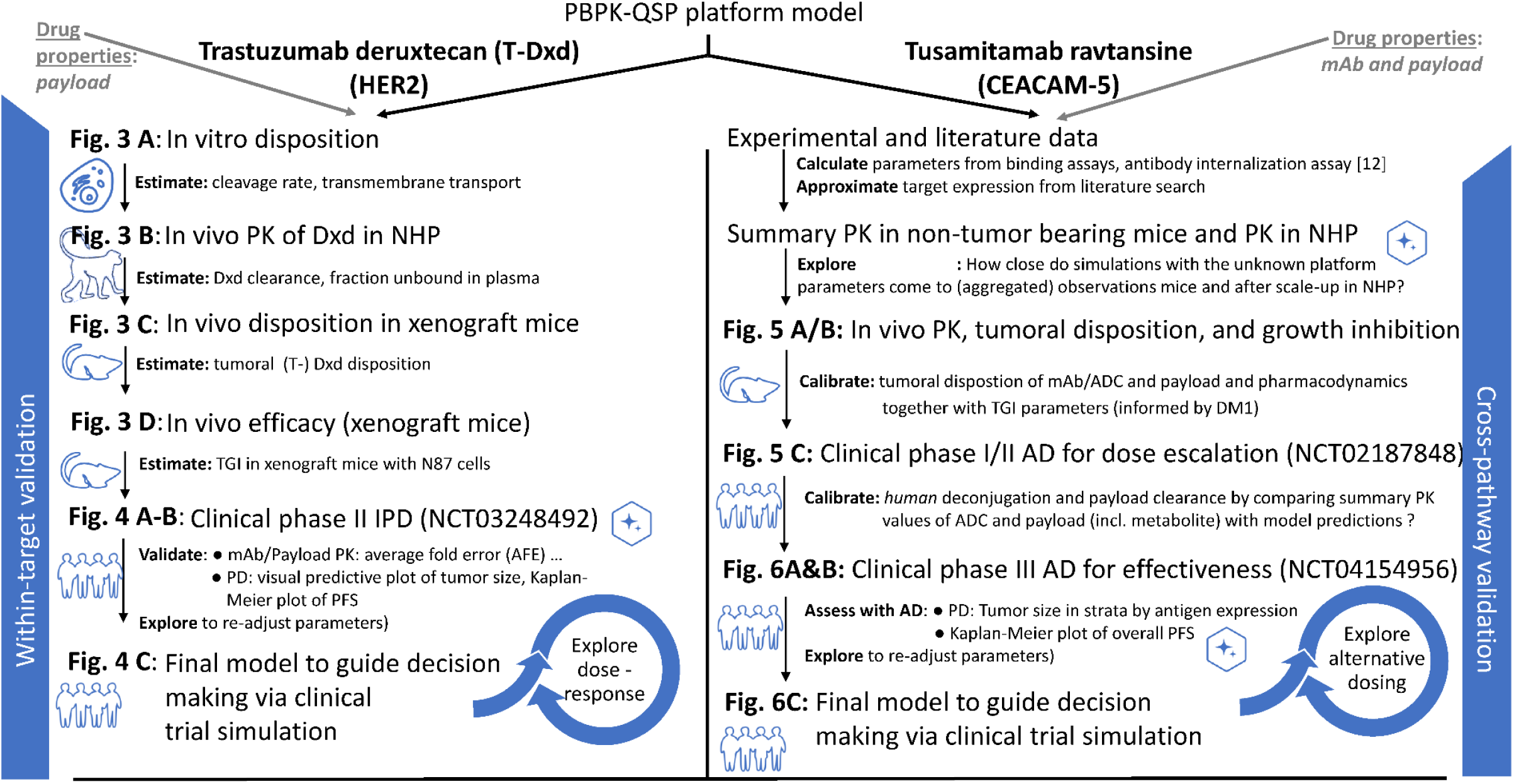
The procedures for within-target validation (left side) and cross-pathway validation (right side) of the platform PBPK-QSP model originally developed for T-DM1. AD: Aggregate data; IPD: Individual patient data; mAb: monoclonal antibody; NHP: Non-human primates; PD: pharmacodynamics; PFS: progression-free survival; PK: pharmacokinetics; SAR408701: Tusamitamab ravtansine; (T-)Dxd: trastuzumab deruxtran;;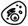: In vitro cell culture experiment; : In vivo mouse experiment; 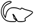: In vivo NHP experiment;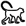: Clinical trial data.

### Developing a T-DM1 PBPK-QSP platform model

#### Model sources for the T-DM1 PBPK-QSP platform model targeting HER2 patients with HER2-positive breast cancer

Existing models for describing the PK and the PK-PD relationship of monoclonal antibodies were adopted from the literature and modified for our particular purpose. In brief, we report the sources and noteworthy adaptations. The starting point was the PBPK platform model for mAbs from Shah and Betts [15]. The physiological parameters in the original PBPK model were fixed with theoretical values from respective species. In line with the translational spirit of a drug development pipeline, we mapped the physiological parameters across different species by allometric scaling as follows:

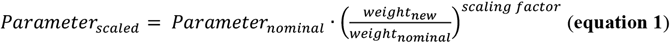

with a default *scaling factor* for distributional rate constants set as 0.75 and for volumes of 1. In particular, the nominal values were taken from the mouse physiology in the PBPK model of Shah and Betts [15] and scaled up as appropriate across species. This PBPK backbone for mAb was extended so that the payload can be released from the antibody part of an ADC, as described for T-DM1 with non-cleavable linkers [16] or with cleavable linkers [17, 18].

For tumoral disposition, we used the Krogh cylinder model [19] and calculated transfer constants for the amount of permeation and diffusion into the tumor extracellular matrix with:

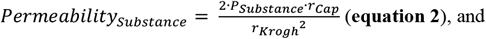

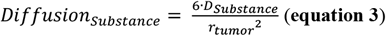

where *r*_*Cap*_ is the radius of tumor blood capillary, *r* _*Krogh*_ is the Krogh cylinder radius for the average distance between two capillaries, *r*_*tumor*_ is the tumor radius, *P* is the permeability coefficient for the respective *Substance* (mAb/ADC or payload) across and around the tumor blood vessels, and *D* is the respective diffusion coefficient. Thus, transfer from the central compartment to the tumor extracellular matrix is driven by a concentration (*C*) gradient Δ*C* = *C*_*central*_ − *C*_*extracellular matrix*_, so that *transfer* = (*Permeability*_*Substance*_ + *Diffusion*_*Substance*_ ) · Δ*C*.

Linking of tumoral payload PK to PD was achieved by the platform model based on empirical PK for T-DM1 and T-Dxd [7] including a translational pipeline study of T-DM1 [8] and the empirical PK-PD in vivo model for T-Dxd [20]. As such, we adopted the natural HER2 (human epidermal growth factor receptor 2) turnover as described by Scheuher et al. [7]. However, for the cellular HER2 trafficking upon binding of an antibody, we used an earlier concept to describe it by a common endosomal processing rate for the recycled and degraded HER2 portion [21]. The tumor growth inhibition (TGI) model is also based on Haddish-Berhane et al. [22], where intra-cellular payload concentration is the driver of tumor killing [23]:

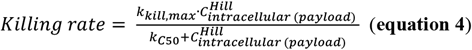

where *k*_*kill,max*_ is the maximum killing rate, *k*_*C50*_ is the concentration for half-maximum killing rate, *C*_*intracellular (payload)*_ is the intracellular payload concentration, and *Hill* is the (optional) Hill coefficient.

A single model coding can be applied for in-vitro, in-vivo preclinical, and clinical purposes (refer to the coding in the Supplementary Information, as it shares a common model structure for various applications). This flexibility is achieved by inactivating certain modules depending on the modeling purpose. For example, when applying the model for in-vitro data analysis, the tumor component for non-tumor-bearing mice can be deactivated (see **Supplementary Figure S1**).

#### Re-calibration of the T-DM1 PBPK-QSP platform model

To re-calibrate our T-DM1 model, we used the same data that have already been used for the development of the original PBPK or QSP model [7, 24]. While adopting most of the parameter values from the original models, certain parameters had to be re-estimated in our PBPK-QSP model. For this purpose, we explored influential parameters by means of sensitivity analysis [25] and particularly re-estimated the parameters of the Krogh cylinder model (**equations 2 and 3**) and the TGI model (**equation 4**). We compared predictions with reconstructed IPD from published Kaplan-Meier curves [26]. At the cellular level, we used the PK data for mAb disposition by Austin et al. [27] and payload disposition by Erickson et al. [28]. Our predictions on in-vivo tumoral PK and TGI were validated with data from Erickson et al. [28] and Haddish-Berhane et al. [22], respectively. On the clinical level, we compared model predictions to the summary PK data from Singh & Shah [8] and progression-free survival (PFS) data from a phase II clinical trial [29].

#### Quantifying translational uncertainty of the T-DM1 PBPK-QSP platform model

Translation approaches can be categorized to forward and backward translations. The typical forward translation begins with collecting data from preclinical experiments (e.g. in vitro cell culture, in vivo xenograft mice) to prospectively predict the drug response in patient populations. Conversely, for backward translation, patient parameters can be determined retrospectively for platform model development using clinical data. Hence, the quantitative discrepancy between parameter estimations derived from forward and backward translation provides insight into the uncertainty inherent in translational approaches from preclinical discovery to clinical settings. From 100 clinical trial simulations (CTS) of T-DM1, we compared the PD parameters including inter-individual variability (IIV) from **equation 4** [7, 8], either estimated based on N87 xenograft mice PK/PD data or retrospectively re-calibrated to better fit the observed efficacy data from a phase II clinical trial [8, 29].

### Model exploration and validation

In terms of model exploration and validation, we simulated PK and PD profiles [30], conducted parameter sensitivity analysis, and assessed the performance of data fittings based on average fold error (AFE), a metric to quantify the logarithmic fold-change of predicted values (*Pred*) to observed values (*Obs*):

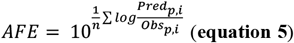

where *n* is the number of observations from patients *p* at times *i*. Moreover, we generated visual predictive plots, e.g., for validating tumor growth inhibition data.

### Application to other ADCs and settings

#### Translational modeling of T-Dxd targeting HER2 in patients with HER2-positive breast cancer (within-target validation)

Figure 1. summarized the workflow and data that was used for model re-calibration and validation. T-Dxd is a HER2-targeting ADC to which a highly potent topoisomerase I inhibitor is conjugated via a cleavable linker in order to exert its cytotoxic effect by damaging DNA [31]. Thus, drug-specific parameters related to the payload and its interaction with the system were adapted in the PBPK-QSP platform being developed for T-DM1, such as binding capacities and the abundance of the intracellular topoisomerase I binding target. Although certain estimated system parameters were published [7, 20], they need to be re-calibrated or re-estimated, due to differences in model structure. The Dxd tumor partition coefficients were calculated according to Poulin and Theil using its physicochemical properties [32]. Permeability surface area values were assumed to be the same as DM1 given the lack of data from permeability experiments.

Using the data from cellular disposition studies [20], the payload deconjugation, payloads’s tumor influx/efflux, target internalization rates, and the fraction of target recycling were re-estimated. Based on the PK data from Dxd administered to non-human primates (NHP) [33], the hepatic clearance and the fraction unbound in plasma were re-estimated. Intra-tumoral disposition data for Dxd [20] were used to re-estimate Dxd distribution and permeability within the Krogh cylinder model (see **equations 2 and 3**). The PD parameters for the TGI model including the drug potency, the maximal tumor killing rate, and the Hill coefficient (**equation 4**) were estimated using TGI data in xenograft mice [31].

Subsequently, the drug-specific parameter values estimated from preclinical data were fixed in the human PBPK-QSP model to validate with clinical IPD data. Having access to phase II T-Dxd data [11] via www.vivli.org, we randomly selected 16 individuals for PK validation and 90 individuals for longitudinal evaluation of tumor size (sum of longest diameter, SLD). Key sensitive parameters identified from a global sensitivity analysis were re-re-calibrated and associated with inter-individual variability (IIV) to fit SLD and time-to-progression Kaplan-Meier profiles. This final model was used to simulate dose-response relationships of objective response rates (ORR) in a simulation consisting of 100 virtual trials with 90 patients each, to reflect the actual clinical trial design.

#### Translational modeling of tusamitamab ravtansine targeting CEACAM5 in patients with lung cancer (cross-pathway validation)

For cross-pathway validation, we adapted our platform to tusamitamab ravtansine, an ADC targeting CEACAM5 with the non-cleavable linker-payload DM4, a structural analog of DM1. This adaptation required to maintain certain payload properties from the T-DM1 platform. We fixed CEACAM5 binding affinity parameter with in-vitro measurement to estimate internalization rates in tumor-bearing mice (for the sequence of re-calibration steps, see also **Figure 1**). CEACAM5 receptor density was approximated based on the literature of non-small cell lung cancer (NSCLC) tissue expression profiles. For the payload DM4, tissue partition coefficients were calculated using the Poulin and Theil approach [32]. Nonetheless, the intricate metabolic profile of DM4 presents a challenge to model the payload profiles. Thus, despite the availability of PK measurements of DM4 and its metabolites (methyl-DM4, and linker-bound forms), we needed to simplify the model by predicting the total payload as a summation of molar quantities of DM4 and its active metabolites (assuming equal potency, respectively). This simplification was deemed acceptable given that all isoforms are equally potent tubulin inhibitors [34, 35]. By assuming the same payload settings that derived from DM1 for DM4, it was possible to assess the predictability of the model against PK data from non-tumor bearing mice and PK data from NHP at the early stage of drug discovery.

In the context of PD modeling, although information on tumor growth dynamics could be sourced from the literature [14], numerous parameters needed to be adapted at this stage. These included cellular influx, distribution and permeability of ADC and its payload, payload clearance, and PD parameters related to TGI. Consequently, this led to over-parameterization, so that the parameters were only be calibrated to fit aggregated and individual PK-PD data from tumor-bearing mice experiments, instead of being estimated using Monolix. Given that the data from lung cell line mice were not available, the TGI data from colorectal cancer xenograft mice models (CR-IGR-034P) were used for the parameter re-calibration. Whereas payload deconjugation rate and clearance parameters were calibrated using clinical PK data of ADC and DM4 PK from phase I/II study (NCT02187848 [13]).

Then, the refined model was validated against aggregated efficacy data from phase III in NSCLC patients (NCT04154956). In particular, model predictions of longitudinal tumor size and Kaplan-Meier time-to-progression were compared with observations, wherein tumor size trajectories were stratified based on patients’ CEACAM5 expression levels, categorized as either low or high. Following the validations, tusamitamab ravtansine dose-response of percentage reduction in tumor size were simulated to explored alternative dosing from existing clinical dosing by assuming that dose-limiting toxicities were primarily driven by plasma ADC *C*_*max*_ [36], while ADC tumor PK exposure (i.e., AUC) and CEACAM5 receptor occupancy were hypothesized to be drivers for efficacy.

### Software

Ordinary differential equations were implemented using the R package rxode2 [37], which also includes a global sensitivity analysis [25]. Parameter estimation was conducted using Monolix (version 2024R1) to fit actual data. To extract observations from plots, we digitized experimental data using WebPlotDigitizer (version 5.1, https://automeris.io).

## Results

### Model platform development using T-DM1 in patients with breast cancer

The PBPK-QSP platform model of T-DM1 was successfully re-calibrated using pre-clinical and clinical PK and PD data. **Supplementary Figure S2** recapitulates the stepwise procedure from (**A**) cellular distribution of antibody and (**B**) payload in vitro, (**C**) in vivo distribution in tumor and growth inhibition in xenograft mice, and (**D**) human PK and PFS data. Using tumor-bearing mice data, the parameters from the tumor disposition model (**equations 2** and **3**) and from the tumor killing function were re-estimated (**equation 4**). For forward translation, these parameters were simply adopted in human translational PBPK-QSP model. This revealed over-optimistic PFS predictions that were stratified by the population’s HER2 expression level (**Figure 2A upper panel**). To effectively perform backward translation, sensitivity analyses showed that parameters related to the ADC tumor penetration (**equation 2**) and tumor killing function (**equation 4**) are equally influential in PFS predictions (**Supplementary Figure S3**). Hence, re-calibration of these parameters of ADC tumor permeability (*P*_*ADC*_) by more than 50% of the original value and maximal tumor killing effect (*k*_*kill,max*_) by less than 10% of original value, and potency of payload on tumor killing (*k*_*C50*_) by less than 40% of original value, resulting in improved PFS predictions against observations for HER2 positive population, nevertheless, the under-predictions of PFS for HER2 normal population remained (**Figure 2A lower panel**).

**Figure 2.**
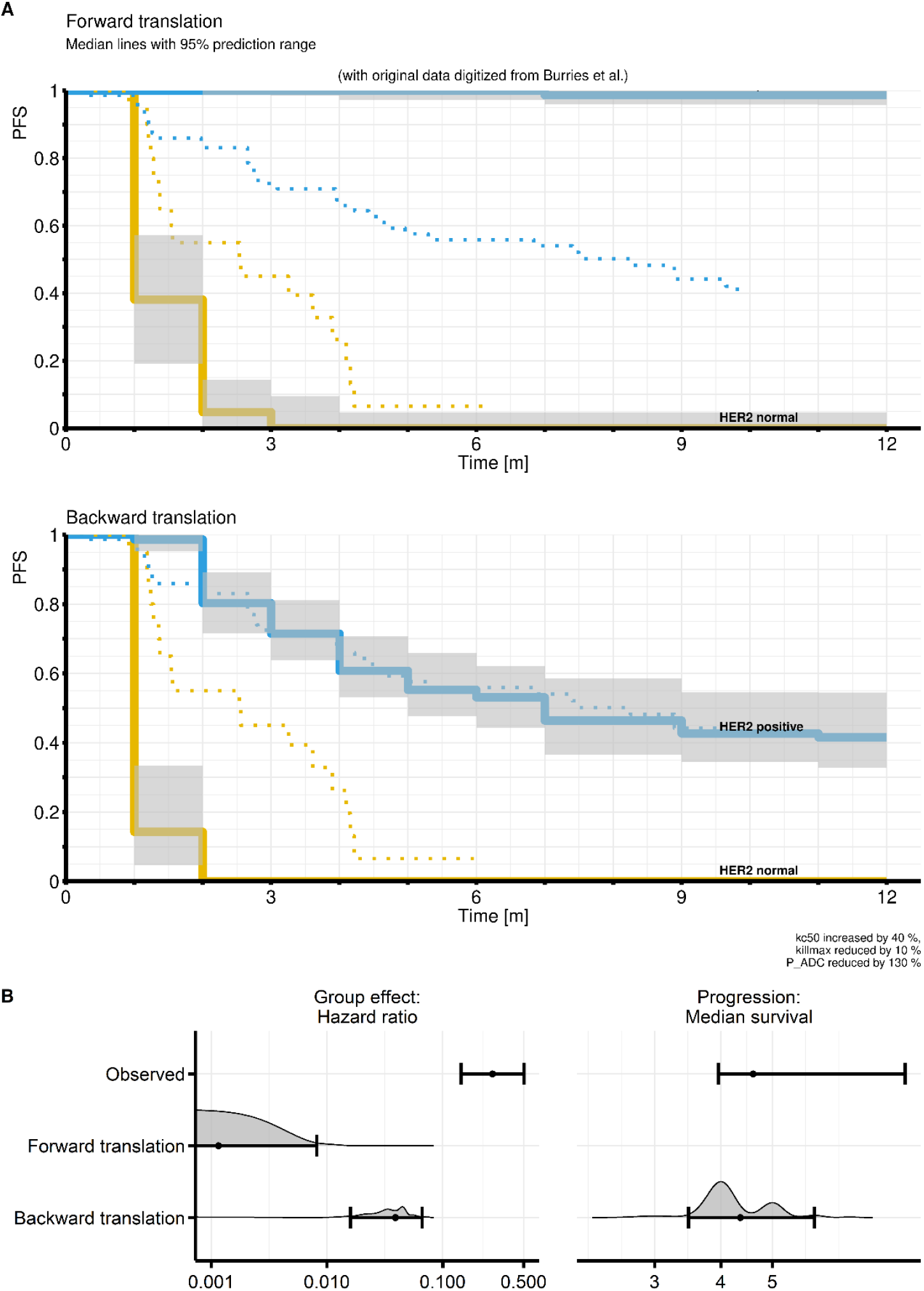
Simulations from the T-DM1 PBPK-QSP model to quantify translational uncertainty by comparison of forward translation and backward translation in 100 clinical trial simulations. (**A**) Kaplan-Meier plots of progression-free survival (PFS) in groups of high (“HER2 positive”, blue line) or normal HER2 (yellow line) expression (median predicted curves and 95 % prediction intervals shown as solid lines on gray areas, dotted lines represent observations from phase II data from Burris et al. [29]). (**B**) Forest plot showing the distribution of hazard ratios for this group comparison (left panel) and the median survival time of both groups in the total cohort (right panel), compared to the observed outcomes phase II trial data [29], respectively.

### Validation of the translational platform model using T-Dxd data in patients with HER2-positive breast cancer

To validate the translational PBPK-QSP platform for the second ADC (i.e., T-Dxd), drug-specific parameters were estimated from in-vitro, NHP, and tumor-bearing mice experiments. Using in-vitro tumor cellular disposition model, the payload (Dxd) cleavage rate was estimated to be 2 · 10^−5^ · *h*^−1^ to adequately fit the Dxd cell-culture media and intracellular concentrations (**Figure 3A**). The Dxd clearance was estimated to be 2-fold higher than DM-1 using the PBPK model to fit plasma Dxd PK data in NHP (**Figure 3B**). With mAb and Dxd collected from plasma and/or tumor in tumor-bearing mice, the rates of tumor diffusion and permeability were estimated to be 9-fold and 6-fold higher than original (DM1) values, respectively, indicating increased Dxd tumor penetration compared to DM1 (**Figure 3C**). Furthermore, Dxd was also found to be more effective in tumor killing than DM1, with respective Dxd parameters being more potent (e.g., 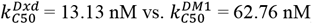) (**Figure 3D)**. Based on our calculation, Dxd can kill about 3-fold faster than DM1 at 50 nM.

**Figure 3.**
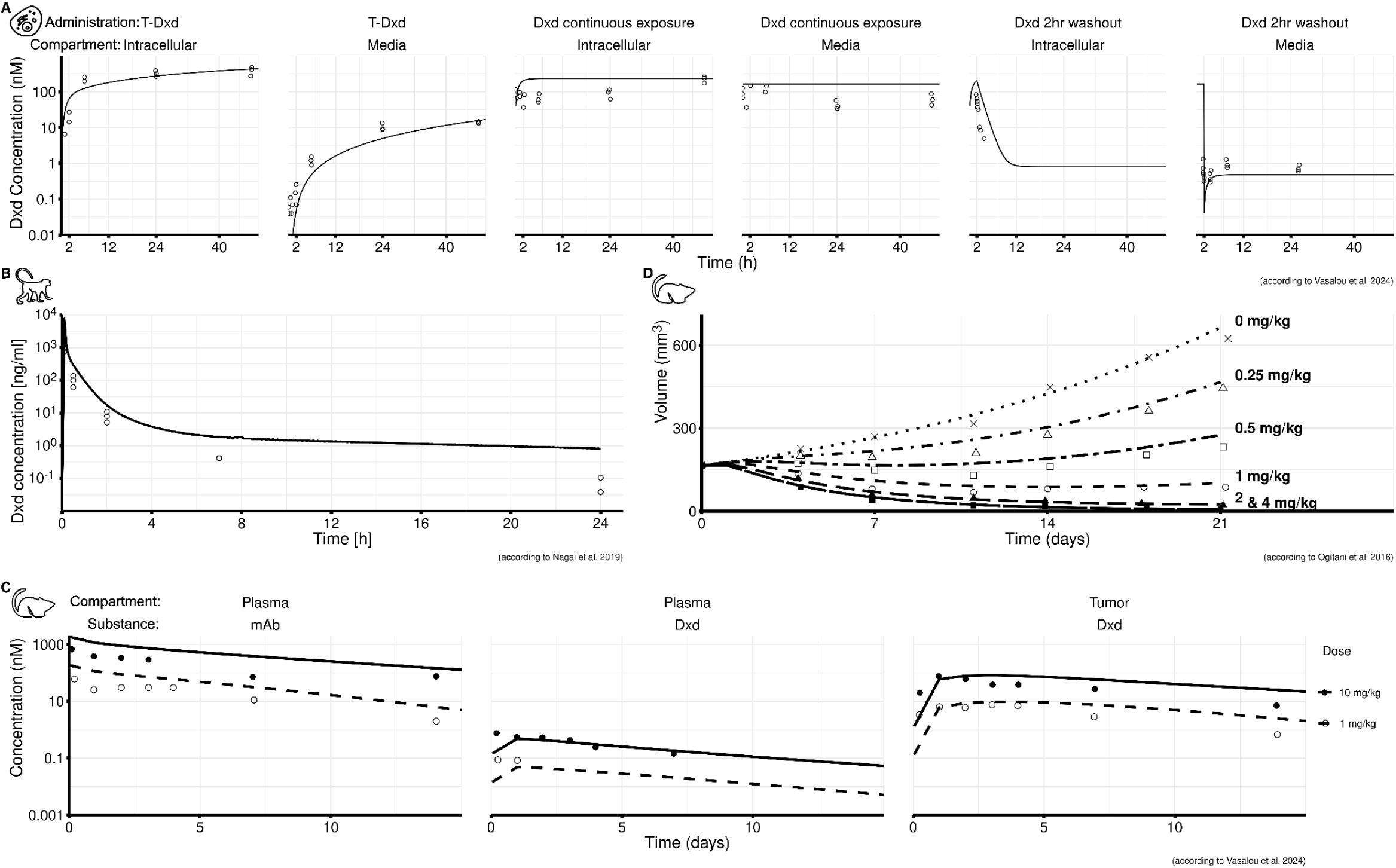
Model fits of T-Dxd against PK (i.e., as total monoclonal antibody, mAb, or Dxd payload) and PD (i.e., tumor volume) data during parameter re-calibration. (**A**) In-vitro (N87 cell line) Dxd profiles measured as intracellular concentration and media concentration following administrations of 97.6 nmol T-Dxd or 162 nmol Dxd to the media (with continuous exposure and washout by refreshing the media after 2h, respectively). (**B**) Dxd plasma PK in NHP following 1 mg/kg Dxd i.v. administration. (**C**) Tumoral disposition in xenograft tumor-bearing mice (NCI-N87) by plasma mAb and Dxd payload PK profiles, and tumoral Dxd PK profiles following 1 and 10 mg/kg single dose i.v. administration of T-Dxd. (**D**) Tumor volume profiles following 0 (control), 0.25, 0.5, 1, 2, and 4 mg/kg single dose i.v. administration of T-Dxd. Lines represent predictions and dots represent observations.

By incorporating drug-specific parameters estimated using pre-clinical data through forward translation, the translational human PBPK-QSP model adequately described plasma mAb PK observations, as prediction error was less than 2-fold (average fold error AFE = 1.45) [**Supplementary Figure S4)**]. Conversely, the Dxd PK was underpredicted (AFE of 0.35), particularly due to the PK measurements collected immediately after T-Dxd administration, which could be explained by immediate payload deconjugation occuring either during or after T-Dxd administration. As a result, *C*_*max*_ was not being captured adequately by the model predictions (**Supplementary Figure S4**). Like T-DM1, the translational PBPK-QSP platform consistently predicted an overly optimistic efficacy, which is demonstrated by a dose-response curve for ORR following forward versus backward translations (**Figure 4C**). By performing backward translation, re-calibrations of tumor killing parameters (*k*_*kill,max*_ from 0.013 to 0.011 h^-1^, Hill’s coefficient from 1.73 to 1.16, *k*_*C*50_ from 13 to 30 nM) and ADC tumor permeability (from 5.28 to 2 *μm* · *h*^−1^) were necessary to match clinical efficacy observations (**Figure 4A & 4B**). As a result, the approved dose of T-Dxd (5.4 mg/kg) approaches the plateau of the dose-response curve (**Figure 4C**), confirming that doses higher than 5.4 mg/kg would not offer additional benefit.

**Figure 4.**
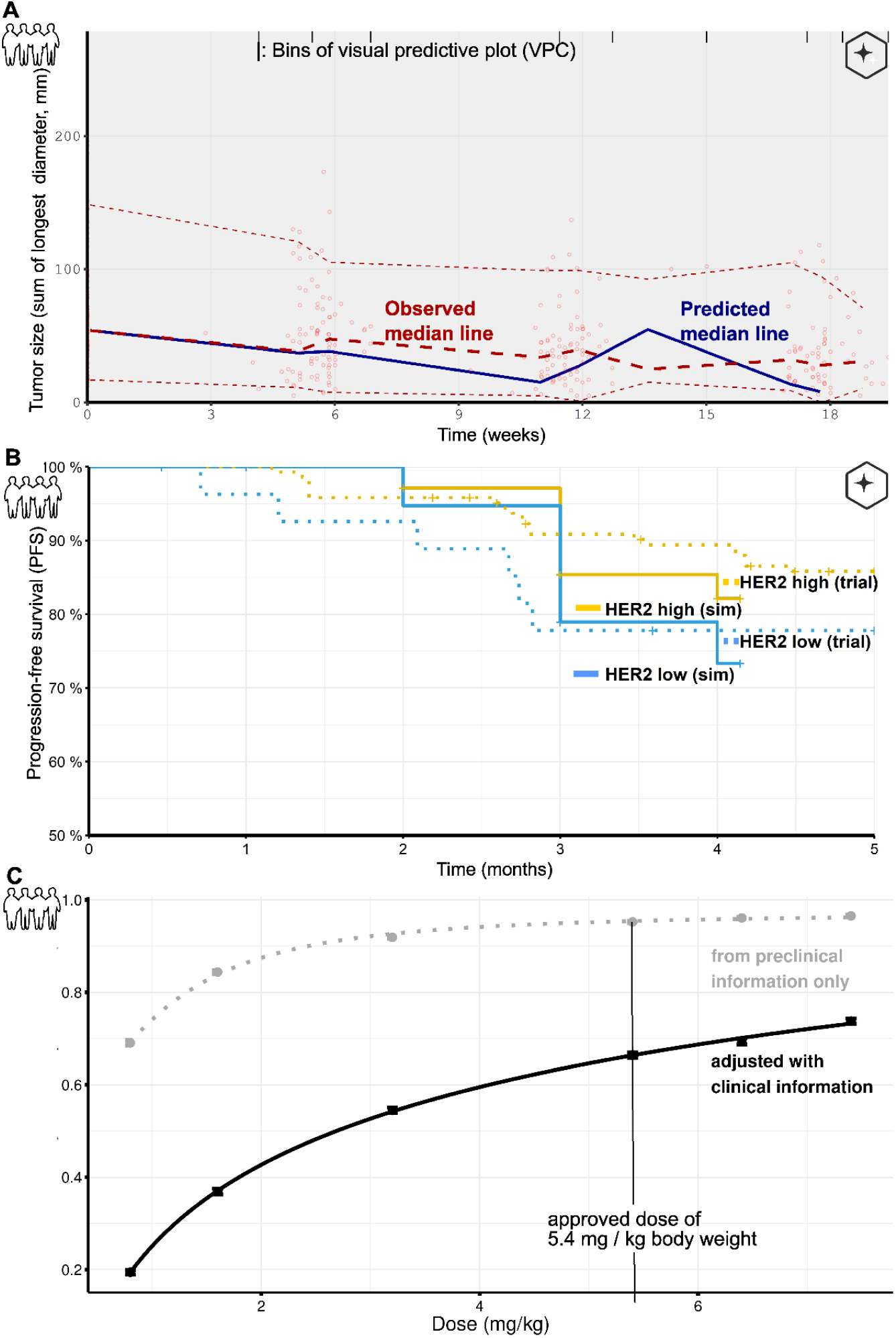
Model fits of T-Dxd against phase II clinical data for PBPK-QSP model validations. (**A**) Visual predictive checks of tumor size (sum of longest diameter; emptied dots represent observations, red dashed lines represent observed median **±** 90% confidence intervals, and blue line represents the predictions). (**B**) Kaplan-Meier plot of PFS by patient populations of low (blue) and high (yellow) HER2 expression (solid lines represent predictions and dotted lines represent observed PFS). (**C**) Simulated population dose-response curve at 0.8, 1.6, 3.2, 5.4, 6.4 and 7.4 mg/kg body weight tri-weekly T-Dxd administration (100 replicates of simulations per treatment group) to record objective response rates (ORR) after one year of follow-up in two scenarios based on preclinical information only (forward translation; grey) and based on re-calibrated parameters (backward translation; black) (vertical line represents approved T-Dxd dose 5.4 mg/kg).

### Validation of the platform model using tusamitamab ravtansine data in NSCLC

To further validate the translational PBPK-QSP model platform, pre-clinical and clinical PK-PD data from the third ADC (i.e., tusamitamab ravtansine targeting CEACAM5 expressed patients with NSCLC) were used to calibrate both disease-specific and drug-specific model parameters for a different tumor-associated antigen and indication as compared to the first two ADCs. Specifically, target expression density of CEACAM5 (5 · 10^5^ receptors per cell) was adapted from literature sources (concentration based on Kim et al. [39] as considered representative for all collected literature sources). The binding affinity constant *K*_*D*_ towards CEACAM5 and the internalization rate constant were fixed to the experimentally determined values 15.04 · 10^−3^ *nM* and to 0.11 · *h*^−1^, respectively. Given the lack of information for CEACAM5 turnover rate, it was assumed to be the same as HER2 receptors. Similarly, parameters related to tumor penetration and payload potency *k*_*C*50_ for tumor killing were re-calibrated to capture the intra-tumoral payload DM4 PK profiles (**Figure 5A**) and TGI (**Figure 5B**) in tumor-bearing mice. During re-calibration process, the parameters of ADC tumor permeability was increased 2-fold, payload potency (*k*_*C*50_) on tumor killing was decreased by approximately 2-fold in relative to (T-)DM1. For forward translation, these re-calibrated parameters were applied in the translational model, however, plasma PK of payload and ADC predictions were partially captured against the observations in patients with NSCLC. Even with re-calibrations of DM4 deconjugation rate constant by 2-fold lower and its clearance by 4-fold lower than DM1, resulting in limited improvement in the model’s fitting of the plasma DM4 and ADC PK data in patients (**Figure 5C**).

**Figure 5.**
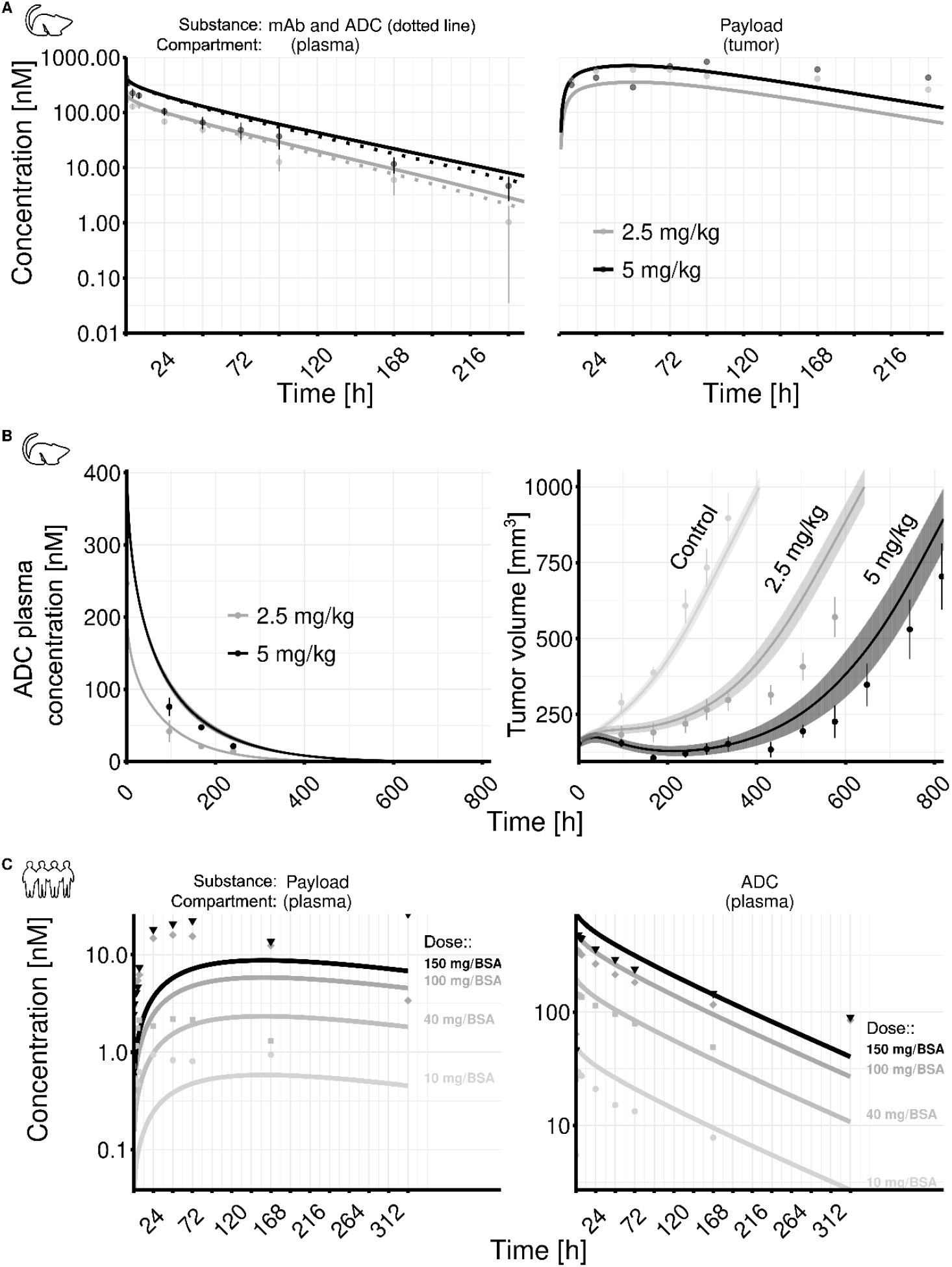
Model fits against tusamitamab ravtansine PK-PD data following model parameters re-calibration (observations represented by dots, predictions represented as solid lines). (**A**) PK of total monoclonal antibody (mAb, solid lines) and intact ADC (dotted lines) upon administration of 2.5 mg/kg (gray) and 5 mg/kg body weight (black) to tumor-bearing mice (patient-derived xenograft). (**B**) Plasma PK of *intact* ADC and corresponding tumor volume profiles following placebo, 2.5 and 5 mg/kg single dose administration to in total 17 tumor-bearing mice (patient-derived xenograft). (**C**) Plasma PK of total payload and intact ADC following 10, 40, 100, and 150 mg/body surface area (BSA) single dose administration from a dose-finding study [13].

We assessed these calibrations by validating with efficacy data in NSCLC patients (see **Supplementary Figure S5A**). Finally, *k*_*C*50_ and *Permeability*_*ADC*_ had to be adjusted by 3-fold, that is far more than expected from the T-DM1 backward translation in order to obtain a congruent tumor size progression (SLD) with loess-fitted aggregated data over three treatment cycles (**Figure 6A**). To better predict late progression after initial response, we successfully added a resistance module to the TGI model, where a certain proportion of tumor cells leaves the pool of proliferating cells (*N1*) with an induction constant *k*_*ind*_ in order to proliferate independently at a lower growth constant than original tumor growth, but eliciting resistant effects to tumor killing (see **Supplementary Material 1**). The extended model improved the PFS predictions when such resistance was randomly introduced to one third of the patients (**Figure 6B**). We continued with this extended model to explore alternative dosing regimens of tusamitamab ravtansine: 75 mg/BSA QW, 75 mg and 50 mg/BSA QW administered alternatively, and fractional 100 mg/BSA Q3W followed by 75 mg/BSA QW, as shown in **Figure 6C**. Simulations showed that alternative dosing regimens yielded similar *C*_*max*_ values, suggesting the dosing could provide comparable safety profile to the original dosing regimen 100 mg/BSA Q2W. In contrast, these regimens predicted a higher CEACAM5 receptor occupancy, potentially enabling greater maximum tumor reduction. Notably, the fractional 100 mg/BSA dose for three weeks, followed by a maintenance dose of 75 mg/BSA QW, could be a reasonable dosing for safety and efficacy, while preserving convenient dosing intervals and providing high average exposure within a short period of fractional dosing.

**Figure 6.**
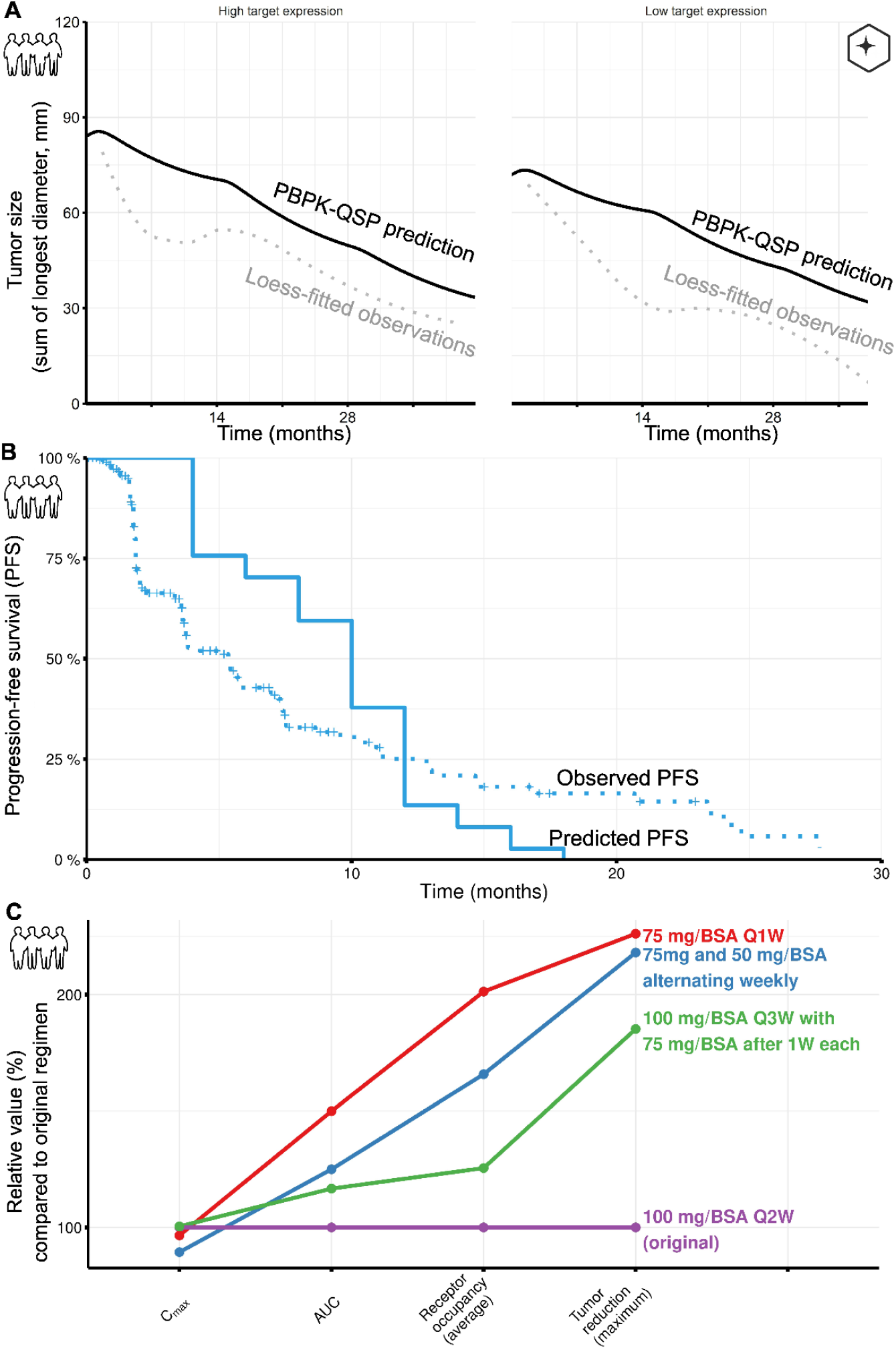
Model fits and simulations from the validated PBPK-QSP model for tusamitamab ravtansine. (**A**) Tumor size PD response after biweekly administration of 100 mg/body surface area in strata of high and low target expression (dotted lines represent a loess fit of observed aggregated data from phase III trial (NCT04154956), solid lines refer to predictions). (**B**) Kaplan-Meier plot of progression-free survival as observed in the tusamitamab ravtansine arm from phase III trial (NCT04154956, dotted line) and the prediction from the model including a tumor resistance module (solid line). (**C**) Clinical trial simulations over six weeks to derive predictions of maximum (peak) plasma concentrations (C_max_), cumulative areas under the concentration time curve (AUC), average percentages in receptor occupancy, and percentage reductions in tumor size, all of which are expressed as percentages related to the originally investigated regimen. The dosing regimens included weekly dosing of 75 mg/BSA (red line), weekly dosing of alternating 75 mg/BSA and 50 mg/BSA (blue line), triweekly dosing of 100 mg/BSA followed by 75 mg/BSA after seven days (green line), and biweekly dosing of 100 mg/BSA.

### Guidance for successful application of a translational modeling platform

Based on the experiences learned from the two cases of translation, we propose general recommendations for applying ADC platform models to new clinical candidates. **Figure 7** represents an Ishikawa-like flowchart [52] outlining essential experimental data, alternatives when experimental data are lacking, and complementary experiments to refine or confirm platform estimates. We categorized the anticipated variability and uncertainty of key parameters while recommending uncertainty assessment to evaluate the impacts of parameter approximations from different sources. This is especially important for forward translation, where we had to correct tumoral disposition and killing capacity by factor two after backward verification in our within-target validation, and up to factor of three in cross-pathway validation (applies the translational approach other than breast cancer). Under the real practical human translation, we thus advise applying such conservative correction factors to avoid overly optimistic predictions on efficacy for mitigating the risk of clinical failure.

**Figure 7.**
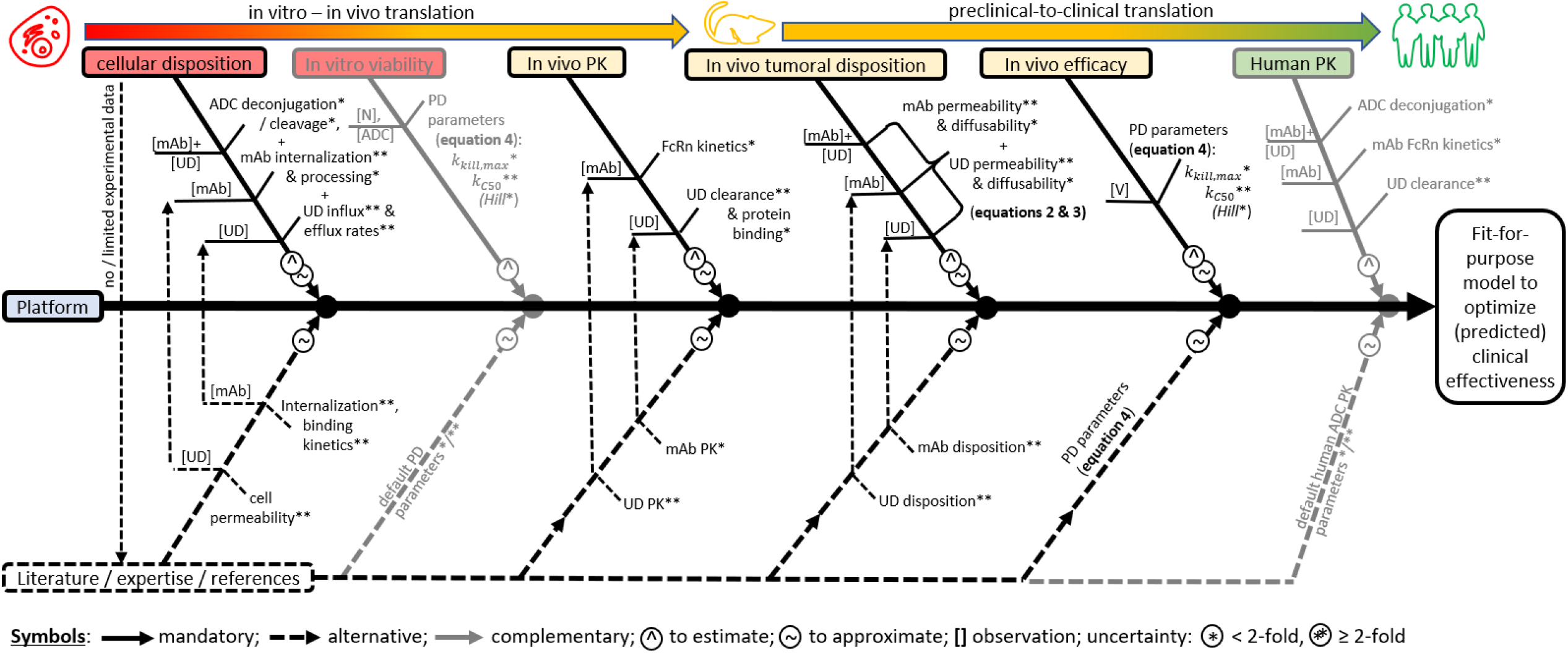
Ishikawa-like flowchart outlining recommended workflow for making a platform model fit-for-purpose allowing to simulate clinical scenarios of interest. The platform model requires re-calibration to the substance of interest, for which dedicated experiments or other source of information for the target variables (left-hand side of the Ishikawa fish bone []) exist that help to estimate or approximate respective model parameters (right-hand side fish bone). The upper part shows a series of experiments that is recommendable to be performed. ADC: antibody-drug conjugate; FcRn: neonatal fragment crystallizable (Fc) receptor; *Hill*: Hill coefficient; *k*_*kill,max*_: maximum killing rate; *k*_*C50*_: concentration for half-maximum killing rate; mAb: monoclonal antibody; N: number of cells; PD: pharmacodynamic(s); PK: pharmacokinetics; UD: unbound drug (payload); V:tumor volume

## Discussion

### Brief synopsis of key findings

We successfully developed and validated a modular PBPK-QSP model for simulating ADCs doses, as demonstrated in breast cancer and NSCLC. By integrating multiple components, our platform model covers applications from in vitro cell culture studies and in vivo PK/PD analyses to patient-specific simulations. This flexibility allows fit-for-purpose modeling and supports informed clinical decision-making. Our case studies, using both original trial data and typical clinical development scenarios with limited information, highlight the complexity of ADC translation and offer a systematic approach for addressing these uncertainties. Considering the challenges and opportunities for dose selection and therapeutic optimization, we propose a recommended workflow to enhance the implementation of translational PBPK-QSP modeling.

### Explanations for key findings

Our ADC PBPK-QSP modeling study revealed important insights into the scalability and behavior of ADC kinetics and pharmacodynamics. Notably, preclinical PK can be well translated to human [15], particularly for IgG monoclonal antibodies that has been confirmed with the two HER2-targeting ADCs: T-DM1 and T-Dxd. Our model also effectively simulated payload kinetics, even with the complexities of multiple payload isoforms formation (i.e. free, metabolites, and linker-bound forms); all of which can introduce significant variability. Exploratory simulations suggested that the initially high observed payload concentrations (particularly for trastuzumab-based ADCs) reflect PK patterns resembling those of direct intravenous administration (data not shown). This underscores the interplay of PK, the payload and the ADC, highlighting the necessity of PBPK modeling to investigate payload release kinetics and its tissue distribution.

Our TGI module also demonstrated robust predictive performance (visually derived using our expert assessment), consistent with previous studies in breast cancer [7, 20, 22, 23] and NSCLC [14]. The ADC and payload tumor penetration emerged as a critical factor influencing therapeutic success and thus underscoring the need for models to accurately capture the tumor PK [38]. While the model can predict long-term efficacy (e.g., PFS) using TGI for various oncology indications [e.g., 7, 8], however, short-term TGI response does not always correlate well with long-term efficacy [40]. Furthermore, the accuracy of the model applying for NSCLC remains highly uncertain due to variable tumor doubling times and complex pathological mechanisms involved, including tumor inhibition resistance and heterogeneity in response to treatment [40-42]. Essentially, a key insight from applying the translational PBPK-QSP model was its tendency to overpredict the TGI effects of ADCs, which may lead to the proposal of lower-than-optimal doses in early clinical trials for novel ADCs. This was reflected by consistently over-optimistic simulations of drug efficacy, as compared to observed clinical outcomes, during model validation using T-Dxd and tusamitamab ravtansine. This can be explained by predicting ADC and payload tumor tissue penetration in patients based on pre-clinical data which remains challenging [43], as there are significant anatomical and physiological differences between human and animals [38, 43-45]. Mice typically have more permeable, vascularized tumors, facilitating ADC accumulation, whereas human tumors are often denser and less accessible, limiting drug delivery [46].

We tested the PBPK-QSP model for dose finding using T-Dxd. Simulations showed that the approved dose of T-Dxd (5.4 mg/kg Q3W [36]) approaches the plateau of the dose-response curve in patients with breast cancer **(Figure 4C)**, indicating that further dose escalation would likely yield minimal additional benefit. These findings reinforce the utility of our PBPK-QSP model as a robust tool for dose finding in the development of novel ADCs. Similarly, using the model to explore alternative dosing regimens for tusamitamab ravtansine were equally satisfactory. Our simulation informs the advantage of selecting adaptive and fractionated dosing strategies based on various metrics, such as *C*_*max*_ and average receptor occupancy. This finding aligns with results from recent case studies with adaptive dosing regimens, particularly improved tolerability and maintained target saturation for gemtuzumab ozogamicin, by applying fractionated dosing scheme, and inotuzumab ozogamicin, by dose adjustment based on patients’ initial responses [36].

### Limitations and Mitigation Strategies

While the PBPK-QSP model was successfully validated with available data, several key limitations were identified. First, while the building blocks for developing the platform model were considered valid, we did not set dedicated tolerance levels for external validation with a different compound (T-Dxd) or even a different pathway (tusamitamab ravtansine). Instead, our conclusions are based on expert judgment after final adjustment (backward translation). This is limiting for the particular application to our examples, but still allows conclusions and lessons-learned, which were our primary goal.

Second, the model did not incorporate critical biological mechanisms, such as HER2 signaling blockade and Fc-effector functions, which may play a significant role in mediating tumor cell killing, particularly for ADCs like T-DM1. With our approach, we aimed to create the most generalized platform possible. For the trastuzumab-based ADCs, the model simulations appeared largely unaffected of the aforementioned potential limitations. However, the simulations for CEACAM5 were less accurate, possibly due to factors like target shedding and expression in various healthy tissues, both of which can complicate PK profiles [6, 7, 12]. While the platform model worked satisfactorily in our validation use-cases, there may be other situations that warrant such modules, because healthy tissues often constitute a larger distribution compartment than tumors potentially leading to substantial target-mediated drug disposition and off-target effects [48].

Third, reliance on a simplified Krogh cylinder model may overestimate ADC tumor penetration. In reality, heterogeneous tumor architecture and diffusional barriers, especially in poorly perfused tumor regions, can significantly hinder deep tissue distribution [45]. To address this, future iterations should integrate more advanced tumor disposition models that explicitly account for diffusional resistance [38, 49, 50].

Fourth, the lack of a direct PD biomarker, such as γH2A.X for DNA double-strand breaks [20], may limit the accuracy of PD effect predictions in early clinical studies [49]. Furthermore, such a marker would have allowed a more direct assessment of when a translation is acceptable. For such continuous biomarkers, it is possible to calculate numeric metrics (e.g., AFE, among many others) that are more concise than, for example, visual assessment of Kaplan-Meier curves (PFS) or visual predictive plots (SLD).

Fifth, omitting the bystander killing effect, a critical mechanism for topoisomerase I inhibitor payloads like DXd, could negatively impact the model’s applicability to tumors with heterogeneous antigen expression [44, 51]. We accounted for this heterogeneity empirically by incorporating inter-individual variability in the killing parameters. This ultimately yielded reliable predictions and consistent results between the trial simulations and actual exposure-response analyses. While this pragmatic approach with our generalized model is effective, it could be further improved mechanistically by extending the model to include the bystander effect.

Many of these limitations are reflected in the flowchart workflow for ADC translation (**Figure 7**). This diagram highlights data gaps, sources of uncertainty, and key decision points, explicitly marking parameters with high expected uncertainty, such as tumor killing parameters and permeability. Despite these challenges, the flowchart outlines a systematic, stepwise approach for developing fit-for-purpose PBPK-QSP model. The process starts with a platform model based on literature, expertise, and reference data. This model is then iteratively being refined by integrating PK and PD data from preclinical and clinical studies, including in vitro cellular disposition, in vivo PK, tumoral disposition, in vivo efficacy, and human PK. At each step, the flowchart distinguishes mandatory, alternative, and complementary data as model inputs either for drug-specific parameter estimation or approximation. This iterative workflow continues whenever new data are available for addressing uncertainties and identifying additional data needed to improve the model predictions, such as the inclusion of bystander effects in the model.

## Conclusions

The preclinical-to-clinical translation of antibody-drug conjugates represents a critical challenge in advancing next-generation therapeutics and optimizing dose-finding strategies. Our work demonstrates how a systematic, quantitative systems pharmacology approach can enhance this translational process through mechanistic understanding and uncertainty quantification. Our platform model use-case and recommended workflow transform these complex mathematical models into an accessible tool for drug developers, enabling faster exploration of alternative dosing regimens and facilitating model-informed decision-making.

## Data Availability

For trastuzumab-based ADCs, this publication is partly based on research data from data contributors Daiichi Sankyo and Roche that has been made available through Vivli, Inc. Vivli has not contributed to or approved, and is not in any way responsible for, the contents of this publication. For cross-pathway validation with SAR408701 (tusamitamab ravtansine), we thankfully acknowledge the provision of data from the ADC development at Sanofi, which are not publicly available, though.

## Supplementary Information

The supplementary material provides additional information.

## Acknowledgments

This research was funded as an award-winning project in the Sanofi iDEA-TECH Awards Europe 2023 entitled “**M**ulti-approach **A**nalysis sta**G**e for model-**I**nformed development of drug-**C**onjugated anti**B**odies with **U**pdates (**L**ive and via **L**iberal software) on **E**xpected **T**reatment outcomes, **MAGIC-BULLET**”.

## Author contributions

All authors were involved in the conception and design of the study, interpretation of data, drafting of the manuscript, and its critical revision. A.D.M. and I.L.-E were additionally involved in original data preparation in the vivli research environment. S.-L.C. and D.V. were additionally involved in original data preparation of SAR408701-related data. ADM developed and applied the models. S.-L.C. and D.V. supervised the project.

## Declarations

### Ethics approval

This publication also used original trial data provided by the data contributors Hoffmann-LaRoche and Daiichi-Sankyo via Vivli Inc. The data request (Vivli ID: 00009229) included an ethical review as standard. Vivli has not contributed to or approved, and is not in any way responsible for, the contents of this publication.

