## Supplementary for "Translational PBPK-QSP modeling platform for antibody-drug conjugates (ADC): within-target and cross-pathway validation to bridge preclinical and clinical results"

*Meid et al.*

### Supplementary Figures

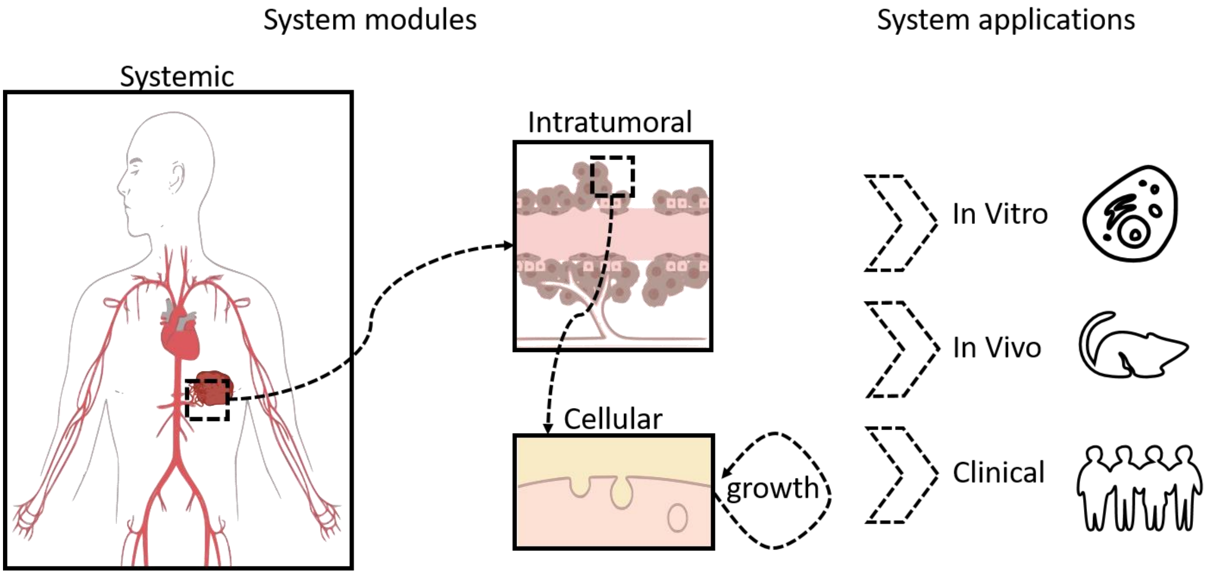


***Figure S1.*** System parts of the PBPK-QSP model, its submodules, and possible applications determined by the choice of model parameters. The model consists of the following **sub-modules**: a PBPK model for antibody and payload kinetics in the systemic circulation, a tumor disposition model for distribution of both into the tumor extracellular environment, a cellular model for antibody binding to its target, internalization, intracellular release of the payload, payload binding to its target, influx and efflux of the tumor cell, and tumor growth (inhibition) model with the drug killing effect elicited by intracellular payload concentration.

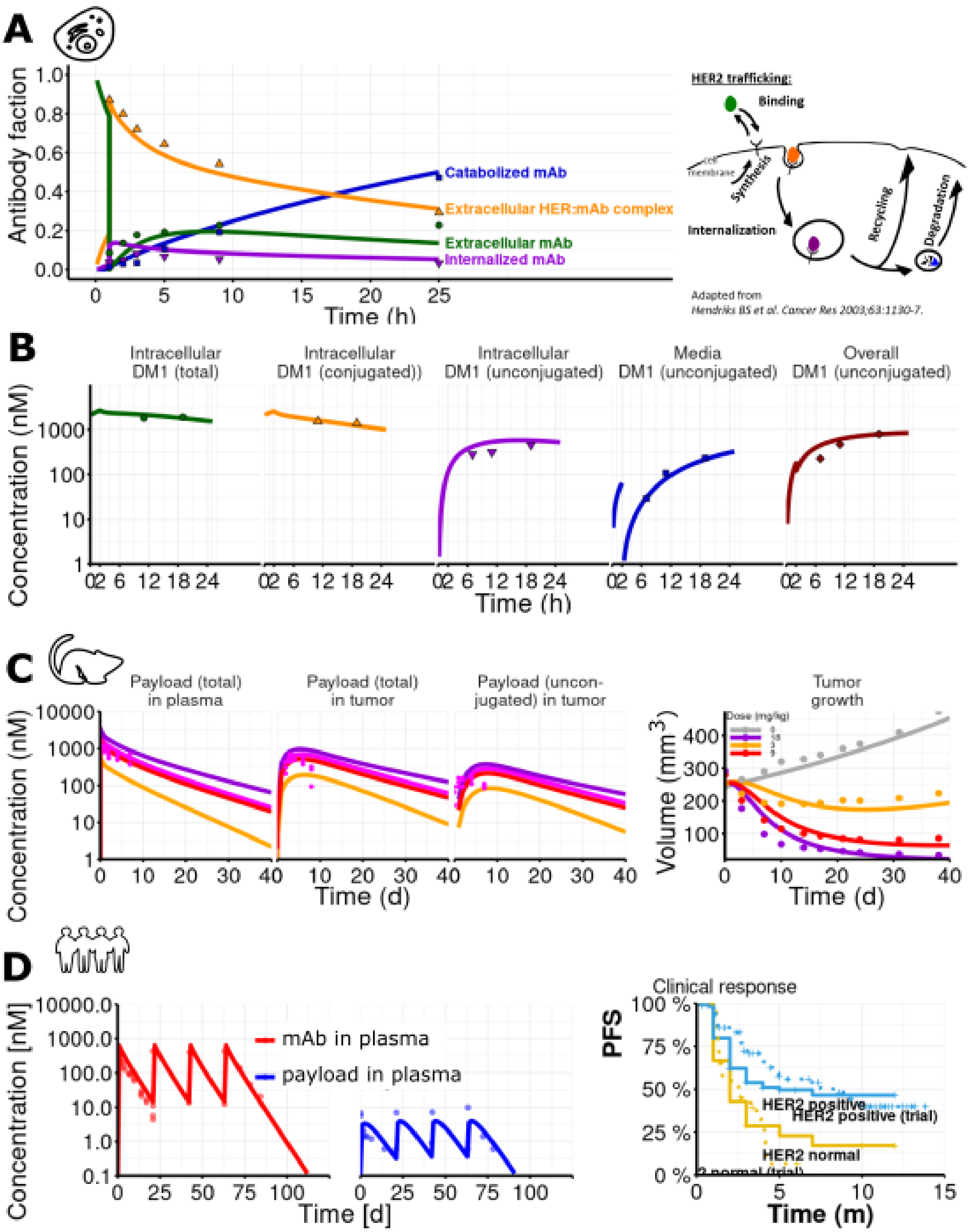


***Figure S2.*** Calibration of the T-DM1 platform model with typical experimental data. (**A**) At the cellular level, antibody trafficking is implemented so that after binding to HER2 and internalization there is a common endosomal processing rate, and then subsequently a portion is recycled and a portion is catabolized (both processes are therefore dependent)[21]. Relative proportions were simulated in these states after a medium exchange after one hour [27]. (**B**) Payload concentrations result from intracellular degradation, among others. Simulations are compared to experimental payload observations in the medium, intracellular, bound to the mAb or free [28]. (**C**) In vivo xenograft studies provided pharmacokinetic data on intra-tumoral disposition (left panel), where simulated pink 12mg/kg line were compared to central and intratumoral observed concentrations [28]. Based on this, several dose-response curves were simulated and assessed for tumor growth inhibition (right panel). (**D**) Human pharmacokinetics were assessed for both for ADC/mAb and free payload drug (left panels). With interindividual variability in key parameters ($k_{kill,max}$, $k_{C50}$,, $P_{ADC}$, tumor doubling time), individual tumor growth over time was predicted for 95 simulated patients according to Burris et al. [29], whereupon progression was derived using RECIST criteria [8]. The dotted lines are based on the phase II clinical trial [29], while the solid lines are model predictions (right panel).

| Parameter | Min | Max |
|---|---|---|
| K_kill_max | 0.00 | 0.01 |
| kc_50 | 10.00 | 100.00 |
| logHill | 0.00 | 0.80 |
| P_ADC | 2.00 | 8.00 |
| DT | 300.00 | 800.00 |
| Kg_Lin | 2587500000.00 | 258750000000.00 |
| tau | 2.00 | 6.00 |

```
[1] "Running sensitivity analysis with 400 simulations, capturing SLD using parameters: K_kill_max, kc_50, logHill, P_ADC, DT, Kg_Lin, tau"
```

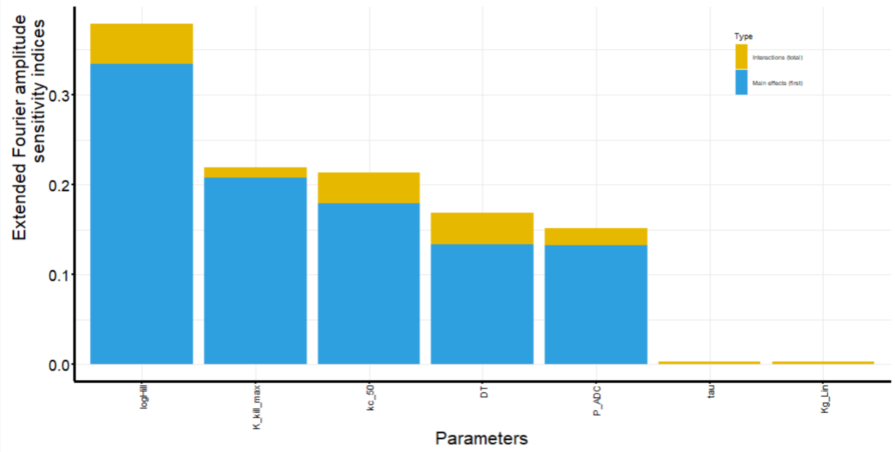


***Figure S3.*** Screenshots of global sensitivity analysis. For T-DM1, the Hill coefficient (operationalized as logHill) was fixed to 1, while T-Dxd it could have particular values.

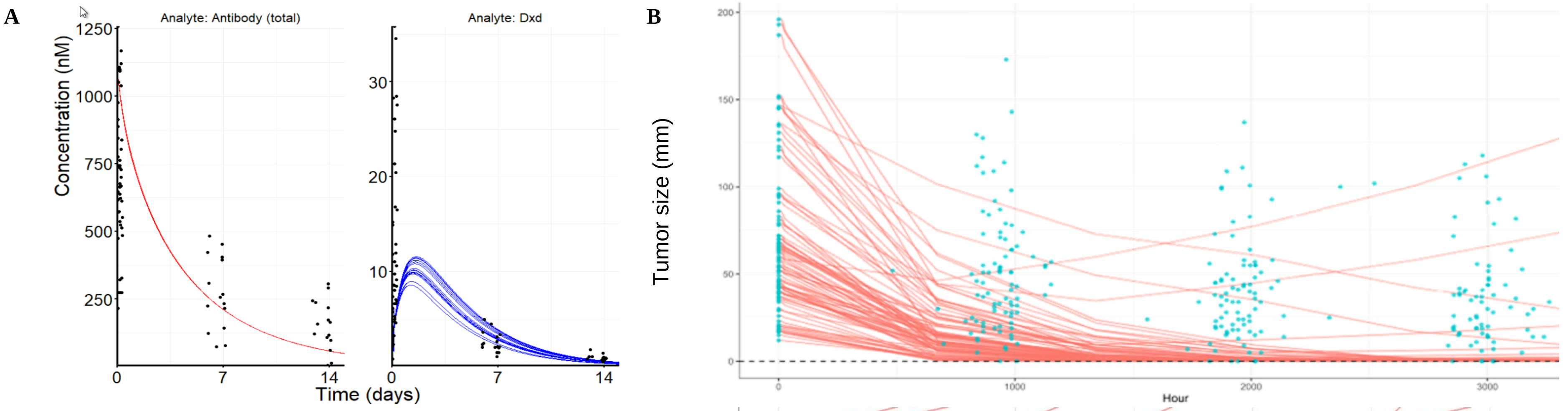


**C** IIV matrix

| | $k_{kill,max}$ | $k_{C50}$ | log Hill coefficient | $Permeability_{ADC}$ | tumor doubling time ($DT$) |
|---|---|---|---|---|---|
| $k_{kill,max}$ | 0.5 | −0.33 | 0.0 | 0.0 | 0.0 |
| $k_{C50}$ | −0.33 | 0.66 | 0.0 | 0.05 | 0.0 |
| log Hill coefficient | 0.0 | 0.0 | 0.0 | 0.0 | 0.0 |
| $Permeability_{ADC}$ | 0.0 | 0.05 | 0.0 | 0.2 | 0.0 |
| tumor doubling time ($DT$) | 0.0 | 0.0 | 0.0 | 0.0 | 0.2 |

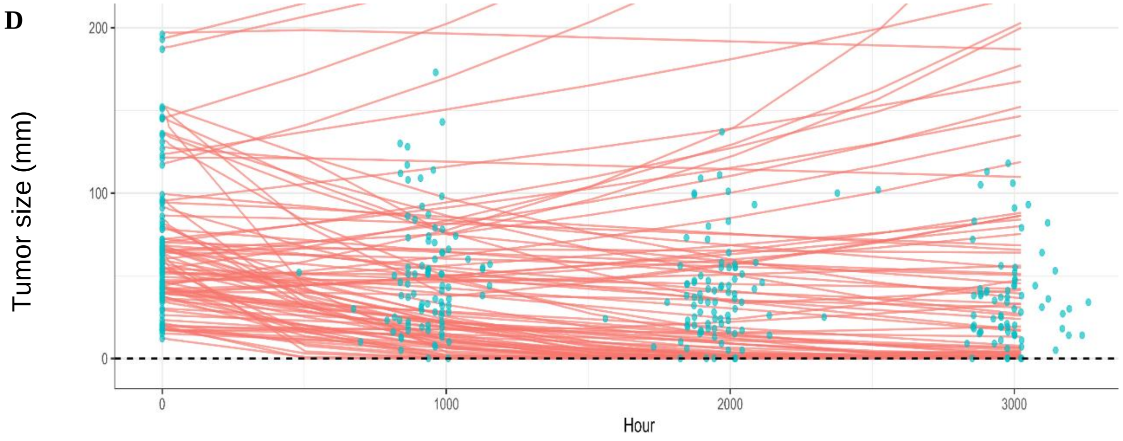


***Figure S4.*** Screenshot of initial validation in of the T-Dxd model with a preclinical parameter. (**A**) Individual observations of plasma concentrations (in nM) of total antibody (shown with a PBPK-simulated typical profile as a red line, left panel) and of unconjugated payload Dxd (shown with simulated profiles representing the randomly chosen 16 patients of various tumor size and HER2 expression level as blue lines, right panel). (**B**) Individual tumor sizes (SLD in mm) measured over time (blue dots) with predicted trajectories for 90 randomly patients of various tumor size and HER2 expression level. (**C**) IIV matrix to update model parameters and (**D**) predictions of individual tumor sizes.

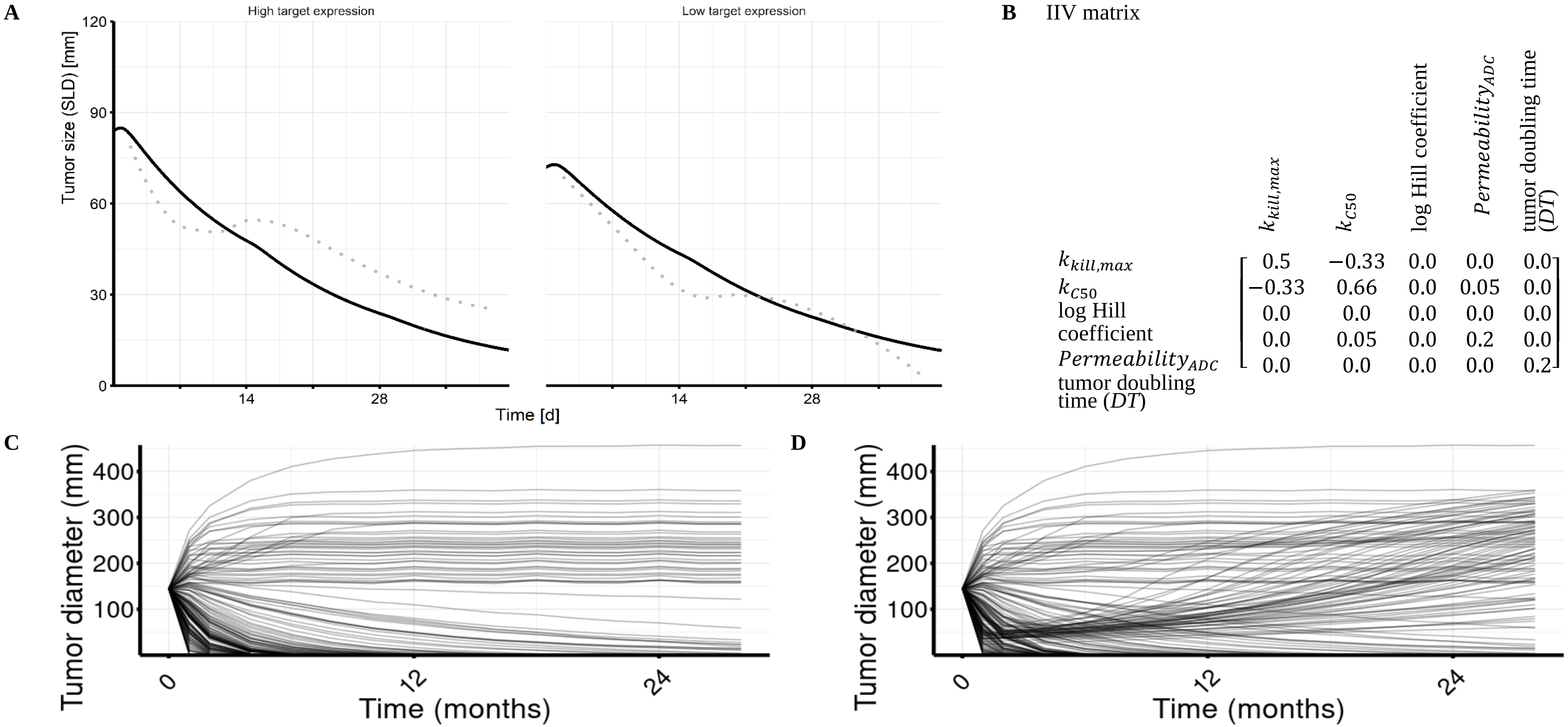


***Figure S5.*** (**A**) Screenshots of initial assessment in of the CEACAM5 model with an unadjusted parameter set. Predicted (black solid lines) and loess-averaged (gray dotted lines) observed trajectories of tumor size with Q2W administration of 100 mg/BSA tusamitamab ravtansine over the first three treatment cycles stratified for patients with high or low CEACAM5 expression. (**B**) Variance-covariance matrix for inter-individual variability in the parameters $k_{kill,max}$, $k_{C50}$, $k_{ind}$, $Permeability_{ADC}$, *and* DT. This matrix is used for clinical trial simulation of 194 patients. This yields individual trajectories of tumor size with Q2W administration of 100 mg/BSA tusamitamab ravtansine (**C**) without resistance or, (**D**) with a resistance module in the model allowing for progression
